# Anti-CMV IgG Titre Determines Organ-Specific Protection Towards Immune Checkpoint Blockade Induced Toxicities

**DOI:** 10.1101/2025.11.07.25339789

**Authors:** Gusztav Milotay, Martin Little, Sophie MacKay, Dylan Muldoon, An-Nhi Huynh, Andrea Farkas, Shawn Sun, Guangyi Niu, Orion Tong, Chelsea A Taylor, Mark R Middleton, Paul Klenerman, Benjamin P Fairfax

## Abstract

Seropositivity for human cytomegalovirus (CMV) is associated with protection against severe (Grade 3+) immune-related adverse events (irAEs) post Immune Checkpoint Blockade (ICB). Here, in a prospectively recruited pan-cancer ICB-treated cohort (n=448 patients), we identify a novel relationship between the relative baseline titre of anti-CMV IgG antibody and organ-specific protection against irAEs. In CMV seropositive patients, whereas anti-CMV IgG antibody level is stable over years, increased pre-treatment titre is independently associated with reduced all-organ Grade 3+ irAEs. This pan-organ association sub-divides into organ-specific effects; protection against non-colitis irAEs being observed only in those with an above median titre of anti-CMV IgG antibody (*P* _High titre_ vs. CMV^−^=2×10^−5^), whereas CMV-related protection against colitis is unrelated to titre (*P* _Low titre_=0.0016, *P* _High titre_=0.0012). We demonstrate that anti-CMV IgG antibody titre is robustly related to peripheral immune subset composition, with higher anti-CMV IgG titre associated with elevated CD4^+^ and CD8^+^ T cell cytotoxicity and effector cell expansion. Conversely, CMV seropositivity is associated with general reduced circulating Tregs irrespective of titre. This work reinforces the importance of CMV in modulating ICB-induced irAEs, revealing a complex relationship between degree of humoral anti-CMV immunity and organ-specific protection, whilst further highlighting the clinical utility of CMV serology in predicting ICB induced irAEs.

## 1 Introduction

Human cytomegalovirus (CMV) is a betaherpes virus that typically causes asymptomatic lifelong latent infection in healthy individuals. Prevalence of CMV infection shows marked variation between countries, with over 80% of people worldwide estimated to be chronically infected, although seroprevalence is lower in many countries including Australia[1], much of Western Europe[2, 3] and the UK, where 56% of assessed white participants were seropositive upon recruitment to the UK Biobank[4]. In health, primary CMV infection is puportedly asymptomatic or leads to non-specific mononucleosis symptoms, whereas *in utero* infection causes primary congenital CMV, the leading viral cause of sensorineural deafness. Similarly, *de novo* CMV infection in immunocompromised individuals may cause life-threatening, multi-organ pathology[5]. Notably, CMV seropositivity is associated with numerous sociodemographic factors including age, sex, non-white ethnicity and Townsend deprivation index[6, 7]. As per other latent herpesviruses, CMV can spontaneously reactivate and, whilst re-activation in healthy individuals is thought to be asymptomatic[8], in immunocompromised individuals it can trigger serious morbidity. Notably, even in health, CMV infection has profound effects on immunity that are most pronounced within the T cell compartment[9]. Specifically, CMV infection, unlike most other viral infections, promotes chronic clonal expansion of effector memory T cells, in a process known as memory inflation[10], which is thought to be due to recurrent re-activation of latent CMV. Memory inflation is therefore a marker of viral latency and has been associated with aging and immunosenescence, although many of the effects of CMV on T cell immunity are distinct to those of aging *per se*[11]. CMV also elicits a humoral response - allowing for serotyping - although the titre of anti-CMV antibody is highly variable between individuals. We have recently demonstrated that IgG seropositivity for CMV is a crucial determinant of oncological outcomes post-Immune Checkpoint Blockade (ICB) treatment in patients with melanoma, as well being associated with the epidemiology of metastatic melanoma[12]. In the same study we observed that CMV^+^ patients have a significantly reduced risk of developing severe (Grade 3+) immune-related adverse events (irAEs), a leading cause of patient morbidity following ICB treatment[13]. Across the OxCITE cohort, a large, prospectively recruited pan-cancer dataset, we note that patients have highly variable CMV IgG titres, as per healthy individuals. The relationship between anti-CMV IgG titre and CMV status in humans is unclear, although high titre is purported to relate to poorer immune control of CMV[10]; whereas in murine CMV infection the antibody titre is related to size of primary inoculate [14]. We have not explored the effect of anti-CMV IgG titre however. To examine this we used the OxCITE study, increasing the sample size used in the analysis of CMV effects from 308 to 448 patients receiving ICB for cancer. We build on our previous observations regarding CMV and ICB-related irAEs to explore the relationship between anti-CMV antibody titre and irAE development. Specifically, we find anti-CMV antibody titre is associated with peripheral immune cell subset composition and provides additional, organ-specific information regarding irAE risk. Thus, in addition to serostatus, anti-CMV antibody titre may serve as a further irAE risk stratification biomarker.

## 2 Results

### 2.1 Anti-CMV titre is stable and independent of covariates

We assessed the clinical and immunological correlates of CMV IgG titre in a pan-cancer cohort from the Oxford Cancer Immunotherapy Toxicity and Efficacy (OxCITE) study (n = 448) receiving either anti-PD-1 alone (sICB) or in combination with anti-CTLA-4 (cICB) as part of standard-of-care treatment. The cohort consisted primarily of melanoma patients (n = 365) as well as non-melanoma cancer subtypes treated by ICB in the absence of chemotherapy or other treatments; namely Mesothelioma (n = 25), Renal Cell Carcinoma (RCC, n = 31), Microsatellite unstable Colorectal cancer (n = 16) and Cutaneous Squamous Cell Carcinoma (cSCC, n = 11). Most treatment was in the metastatic/unresectable setting (n = 387), with the remainder of patients receiving ICB with adjuvant intent (n = 61).

The relationship between pre-treatment CMV IgG titre and clinical covariates was examined. In the CMV^+^ cohort (n = 207) we observed a weak relationship between titre and age at onset of systemic treatment (*rho* = 0.14, *P* = 0.05), supporting a link between the titre of anti-CMV IgG and duration of infection (Ext. Fig.1a)[15]. We did not observe associations between anti-CMV IgG titre and other covariates including biological sex, tumour type and treatment regimens (Ext. Fig.1b-d). To further address whether anti-CMV IgG titre remains stable over time, we selected a cohort of CMV^+^ patients (n = 12) who responded to ICB for at least two years and compared their post-treatment to pre-treatment titres. Consistent with observations from other groups[8, 16], a significant change in titre was not detected over time (*P* = 0.68, Wilcoxon signedrank test), reinforcing a limited coupling between serum antibody levels and time since primary infection (Ext. Fig.1e). Given CMV IgG titres were not normally distributed (Ext. Fig.1f), the seropositive cohort was divided into high and low titre groups, based on the median value (89.4 a.u.ml^−1^, 178.8 a.u.ml^−1^ in undiluted plasma) and such divisions were used for all downstream analysis (n = 241 CMV^−^, n = 103 CMV^+^_Low titre_, n = 104 CMV^+^_High titre_).

### 2.2 CMV titre alters organ specificity of severe irAEs

Revisiting our previous observation of an association between CMV seropositivity and reduced risk of Grade 3+ irAEs in melanoma in this larger pan-cancer cohort, we again observed CMV seropositivity to be associated with significantly reduced risk of all Grade 3+ irAEs (*P* = 2.5 x 10^−4^) (Fig.1a). We proceeded to further assess the influence of IgG titre on severe toxicities using a univariate semi-competing risks model where death was a competing risk[17]. Using this approach, we found that CMV^+^_High titre_ patients were significantly less likely to develop Grade 3+ irAEs compared to CMV^+^_Low titre_ (*P* = 0.0053) and CMV seronegative (*P* = 2.0 x 10^−5^) counterparts (Fig.1b). Conversely, across all irAEs, CMV^+^_Low titre_ patients did not show a significant reduction in Grade 3+ irAEs compared to CMV seronegative patients, although this smaller sample size greatly reduced power and a trend towards protection was observed (*P* = 0.19). Nonetheless, this analysis demonstrates that, in addition to CMV serostatus, the titre of anti-CMV antibody forms a further core determinant of irAE development. As per our previous observation of absence of CMV related-protection against low-grade irAEs[12], we similarly found no influence of anti-CMV IgG titre on low-grade irAEs, reinforcing that the relationship between CMV and irAEs is restricted to high-grade toxicities (Ext. Fig.1g).

**Figure 1:**
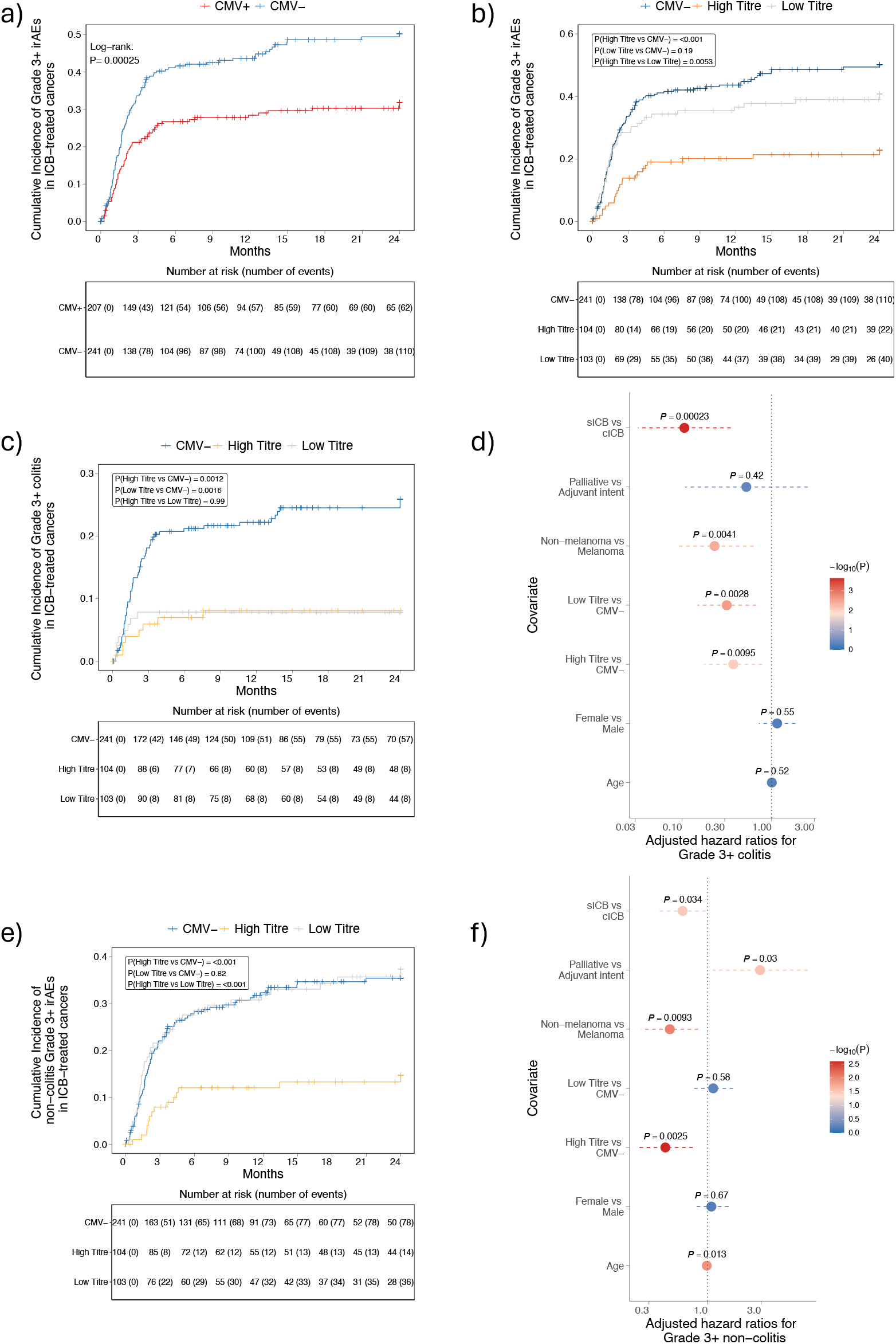
CMV titre is associated with diverse organ-specific risk of severe irAEs. **(a)** Cumulative incidence of developing Grade 3+ irAEs across ICB-treated patients is significantly reduced in CMV^+^ patients 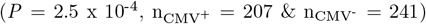. **(b)** Same analysis as a. indicating that risk of Grade 3+ irAEs is reduced in a CMV IgG titre-dependent fashion (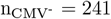, n_Low titre_ = 103, n_High titre_ = 104, two-sided log-rank test). **(c)** Semi-competing risks analysis of Grade 3+ colitis suggests that both CMV^+^_High titre_ and CMV^+^_Low titre_ patients have reduced risk compared to CMV^−^ patients (two-sided logrank test).**(d)**. Multi-variable semi-competing risk analysis, where death is a semi-competing risk highlights that both CMV^+^_High titre_ and CMV^+^_Low titre_ individuals are protected against severe colitis when adjusting for treatment regimen, tumour type, age, and sex (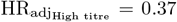, *P* _High titre_ = 0.0095; 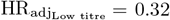, *P* _Low titre_ = 0.0028, Wald test; patients as in a.) **(e)** Same analysis as d. for non-colitis Grade 3+ irAEs. **(f)** Same analysis as c. for severe irAEs excluding colitis indicates that CMV^+^_High titre_ individuals are protected against these toxicities (HR_adj_ = 0.42, *P* = 0.0025).

We previously noted that CMV protection against irAEs showed organ-specificity, with protection against colitis, the most common severe side-effect observed in the OxCITE cohort, being strongest (OR vs. CMV^−^ = 0.39). Notably in this current analysis we find that CMV protection against colitis is independent of anti-CMV IgG titre, with both CMV^+^_High titre_ and CMV^+^_Low titre_ groups demonstrating reduced risk of Grade 3+ colitis compared to CMV^−^ patients (*P* = 0.0012 and *P* = 0.0016, respectively), whilst effect of titre was nonsignificant (*P* = 0.99), indicative of pan-CMV protection in the gut (Fig.1c). A similar result was observed in a multivariable analysis controlling for age, biological sex, tumour type, ICB regimen and treatment intent, with both CMV^+^_High titre_ and CMV^+^_Low titre_ patients being protected against colitis compared to CMV^−^ patients (Fig.1d, P_High titre_ = 0.0095, P_Low titre_ = 0.0028). The other significant covariates associated with colitis being ICB regimen, with cICB demonstrating the known increased risk of irAEs[18]; and cancer type, with non-melanoma patients having reduced risk of severe irAEs. This latter observation, at least in part, likely reflects the switched 1mg:3mg kg^−1^ dose of Ipilimumab:Nivolumab in Mesothelioma and RCC treatment compared to 3mg:1mg kg^−1^ of Ipilimumab:Nivolumab for MM (Supp. Tab.1).

Given the discrepancy between Grade 3+ irAEs and colitis, we next investigated risk of non-colitis Grade 3+ toxicities. Strikingly, we find that contrary to colitis, only CMV^+^_High titre_ patients demonstrated reduced cumulative incidence of Grade 3+ irAEs in other organs compared to both CMV^+^_Low titre_ and CMV seronegative patients (*P* = 5.3 x 10^−4^ and *P* = 3.1 x 10^−4^, respectively) (Fig.1e). Meanwhile, no difference was observed between CMV^+^_Low titre_ and CMV seronegative patients (*P* = 0.82), suggesting that a higher anti-CMV IgG titre is associated with organ-independent control of inflammation. Again, this observation was confirmed using multivariable analysis where CMV^+^_High titre_ individuals were protected against non-colitis Grade 3+ irAEs (*P* = 0.0025, Fig.1f) whereas no effect was noted for the CMV^+^_Low titre_ group (*P* = 0.58). Finally, we explored using a time-independent sensitivity analysis for the most informative threshold of anti-CMV IgG titre for occurence of non-colitis Grade3+ irAEs within a 24-month window of commencing ICB, identifying the cut-off as 95.2a.u.ml^−1^ (190.4a.u.ml^−1^ in undiluted plasma). Given this was almost similar to the median of 89.4a.u.ml^−1^, we proceeded to use this median value in all further analyses.

### 2.3 High CMV titre protects against severe irAEs across treatment types

Grade 3+ irAEs affect *>*60% of patients receiving cICB compared to just 10-15% receiving sICB in clinical trials, highlighting that Ipilimumab is the major driver of severe toxicities[18, 19]. Nevertheless, some organs such as the lungs appear more sensitive to PD-1 blockade, resulting in severe pneumonitis[20]. With this in mind, we sought to identify whether CMV^+^_High titre_ patients were protected from Grade 3+ irAEs in both treatment groups or whether this interaction was specific to CTLA-4 blockade. In the cICB cohort, we found that CMV^+^_High titre_ patients had a reduced cumulative incidence of severe toxicities compared to CMV seronegative individuals 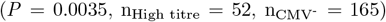 but not compared to CMV^+^_Low titre_ individuals (*P* = 0.061, n_Low titre_ = 61), although the direction of effect was consistent with the complete cohort (Fig.2a). Interestingly, despite high-grade toxicities being rare following sICB treatment and the proportion of patients receiving this treatment being notably lower, we still found that CMV^+^_High titre_ patients had a trend towards lower risk compared to their CMV seronegative counterparts 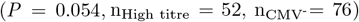 indicating that the interaction with anti-CMV IgG titre is not restricted to Ipilimumab-induced adverse events (Fig.2b).

**Figure 2:**
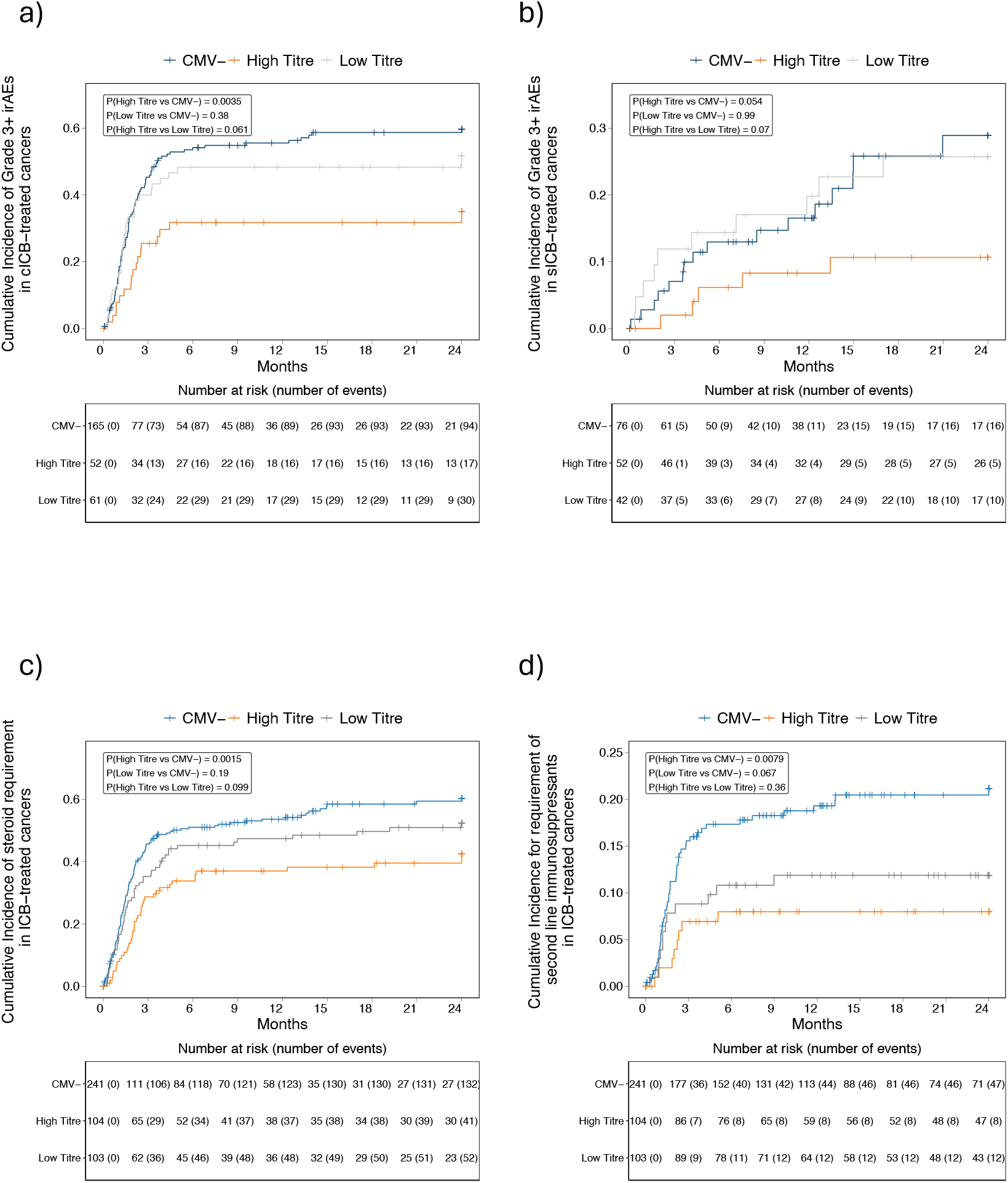
Grade 3+ irAEs according to treatment type and impact on toxicity management. **(a)** Cumulative incidence of developing Grade 3+ irAEs in cICB-treated patients according to CMV titre (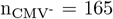, n_Low titre_ = 61, n_High titre_ = 52, two-sided log-rank test). **(b)** As a. for sICB-treated patients (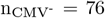, n_Low titre_ = 42, n_High titre_ = 52), highlighting that CMV^+^_High titre_ individuals trend towards reduced risk of Grade 3+ irAEs relative to CMV^−^ patients (*P* = 0.054). Cumulative incidence of steroid requirement **(c)** and second line immunosuppression **(d)** in ICB-treated patients according to CMV IgG titre (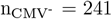, n_Low titre_ = 103, n_High titre_ = 104, two-sided log-rank test).

### 2.4 CMV serostatus but not IgG titre are associated immunosuppressant requirements to manage irAEs

Severe irAEs are typically treated using high dose systemic corticosteroids in the first line, an effect that is increasingly recognised to abate the anti-cancer effects of ICB in meta-analyses[21, 22]. To assess whether titre influenced steroid requirement following an irAE, we performed a semi-competing risks analysis between CMV IgG titre and steroid use, the competing risk being death; finding that although CMV^+^_High titre_ individuals had reduced steroid requirement compared to CMV seronegative patients (*P* = 0.0015) no significant difference was observed between CMV^+^_High titre_ and CMV^+^_Low titre_ patients (*P* = 0.099) (Fig.2c). We also explored the use of second-line immunosuppressants given in the context of steroid failure to control irAEs, finding that there is a significantly reduced requirement for second-line immunosuppression in CMV^+^ patients, although this showed similar direction of effect across both the CMV^+^_High titre_ patients 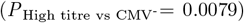 and CMV^+^_Low titre_ patients 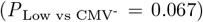, with no discernible difference within CMV^+^ patients (*P* _High vs Low_ = 0.36) (Fig.2d). In summary, this data suggests that irAE management is strongly associated with CMV serostatus, with CMV seropositive patients less likely to require both steroids and second-line immunosuppressants, independent of anti-CMV IgG titre. In part, this observation likely reflects the absence of association between anti-CMV IgG titre and development of colitis, which is the irAE most likely to require both steroids and second-line immunosuppression.

### 2.5 Immunological correlates of CMV titre

Given the divergent irAE profiles within the CMV^+^ cohort according to IgG titre, we sought to identify immunological correlates of anti-CMV IgG titre in the T cell compartment. First, we explored CD8^+^ T cell RNA-sequencing data from baseline, pre-treatment CMV^+^_Low titre_ (n = 57) and CMV^−^ patients (n = 145), identifying 3007 differentially expressed genes (DEGs) (Fig.3a, Supp. Tab.2). Interestingly, when comparing CMV^+^_High titre_ (n = 59) with the same CMV^−^ cohort, we found a significantly larger set of 7,124 DEGs (Fig.3b, Supp. Tab.3), indicating that an elevated anti-CMV IgG titre is associated with magnified transcriptomic differences between CMV^+^ and CMV^−^ patients in the T cell compartment. Among the DEGs we found key markers of cytotoxicity and tissue residency that demonstrated further induction in CMV^+^_High titre_ relative to CMV^+^_Low titre_ patients. To explore these pathways further, we determined the normalised expression of *IFNG*, a marker of cytotoxicity, *KLRD1*, involved in HLA-E recognition by NK and effector T cells, *TBX21*, a key transcription factor for type I T cell differentiation, and *ZNF683* which is a driver of tissue residency[23–26]. All these genes have been reported to play essential roles in shaping both anti-viral and anti-tumour responses[23–26]. Across these markers we found that CMV^+^_High titre_ patients had elevated expression relative to both CMV^+^_Low titre_ and CMV^−^ individuals, most notably for *IFNG* (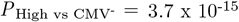, *P* _High vs Low_ = 2.6 x 10^−5^, Fig.4c), suggesting that there is further expansion of effector cells in the T cell compartment of these patients.

**Figure 3:**
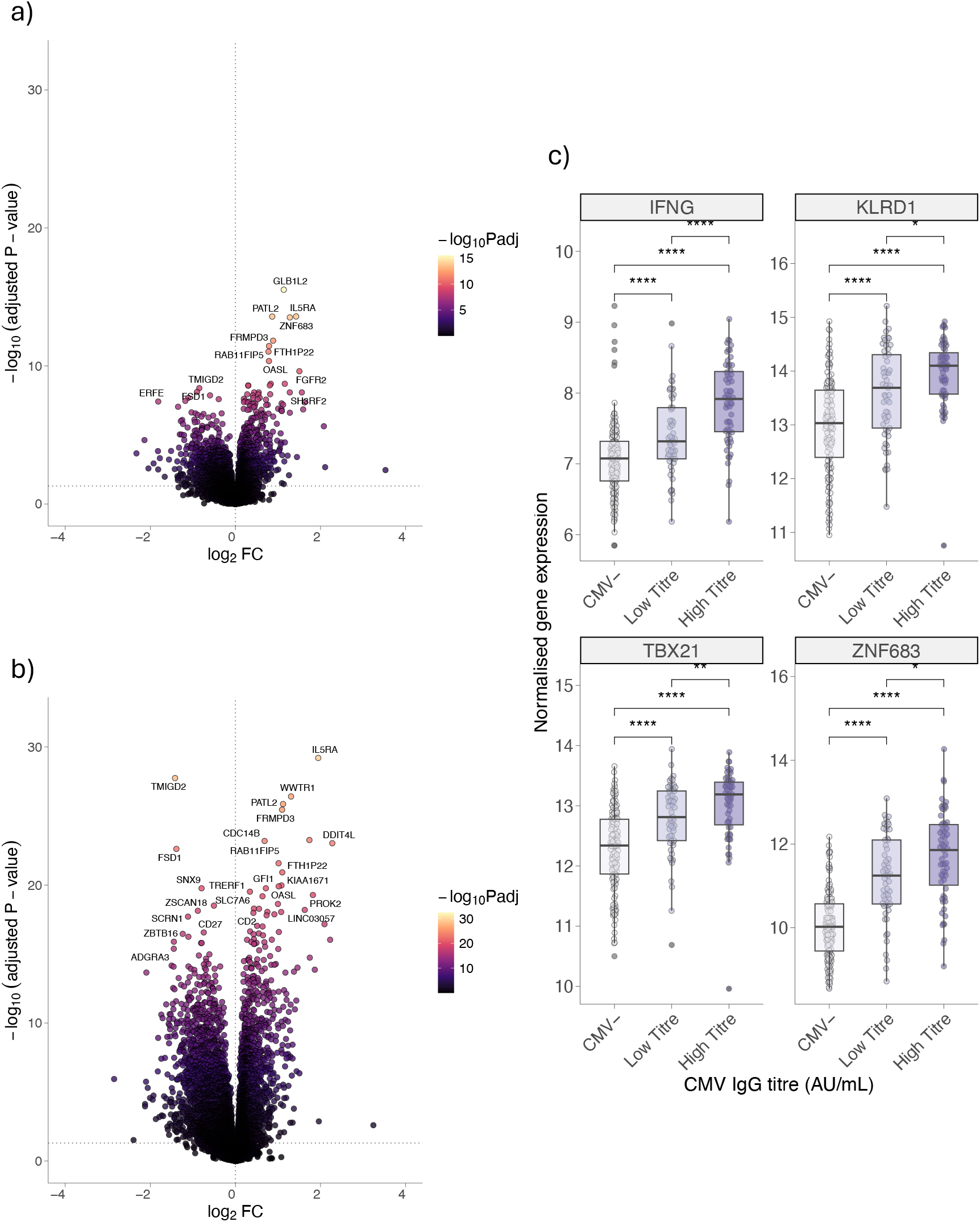
Transcriptomic correlates of CMV titre. **(a)** Differentially expressed genes (DEGs) between CMV^+^_Low titre_ and CMV^−^ patients from pre-treatment CD8^+^ T cells; y axis shows BenjaminiHochberg-corrected-log_10_(Padj) derived from negative binomial Wald test using CMV^−^ samples as a reference (n_Low titre_ = 57, 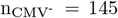). **(b)** As a. assessing differential gene expression between CMV^+^_High titre_ and CMV-patients (n_High titre_ = 59, 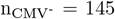). **(c)** Normalised *IFNG, KLRD1, TBX21* and *ZNF683* expression increases in a dose-dependent manner from CMV^−^ to CMV^+^_Low titre_ to CMV^+^_High titre_ patients (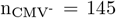, n_Low titre_ = 57, n_High titre_ = 59; *P* values derived from Kruskal-Wallis and post-hoc Dunn tests). Lower and upper box hinges represent the 25th to 75th percentiles, the central line represents the median and whiskers extend to the highest and lowest values no greater than 1.5×IQR.

**Figure 4:**
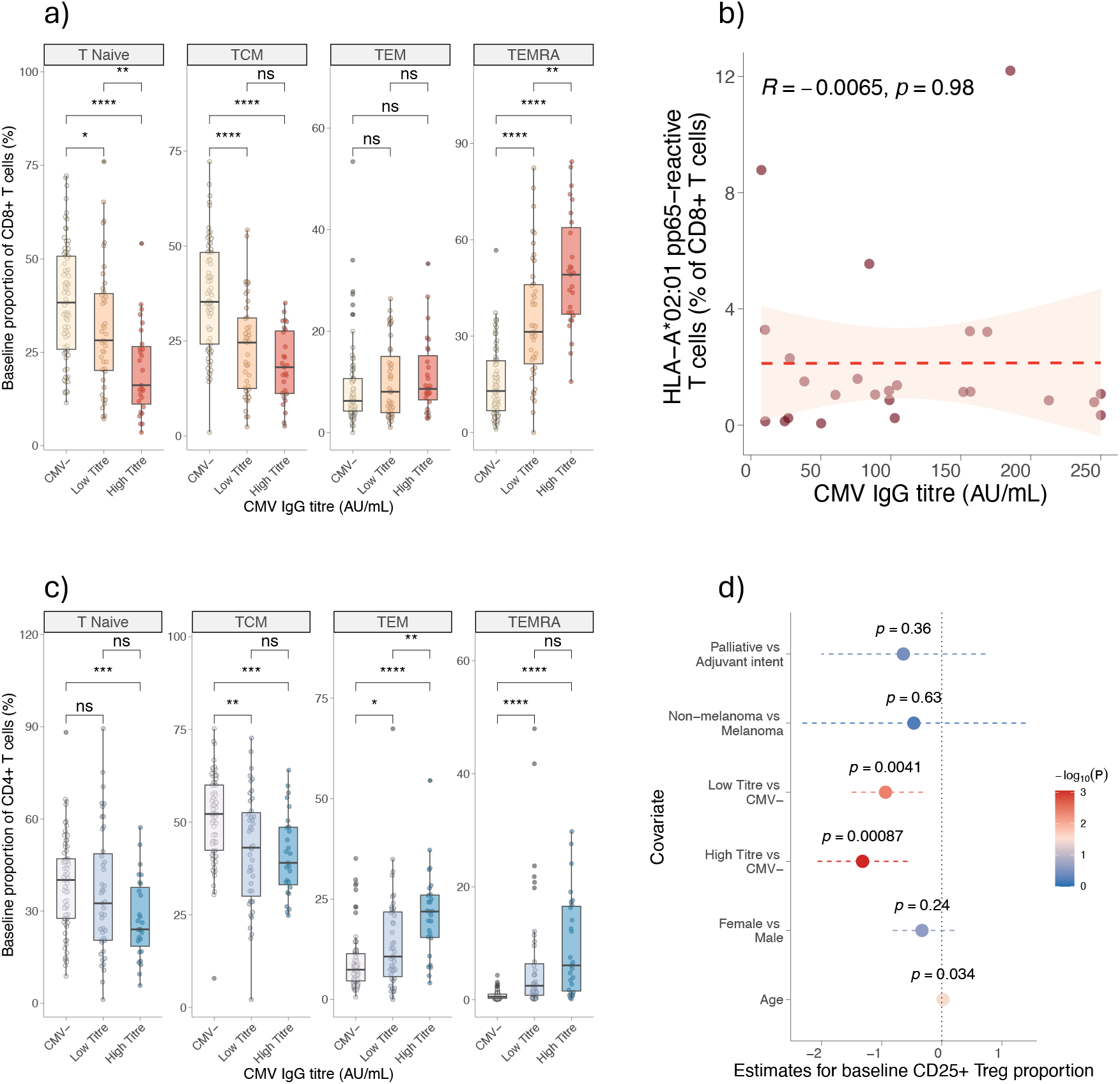
T cell subsets according to CMV titre. **(a)** Flow cytometry-derived pre-treatment CD8^+^ T cell subsets flow cytometry according to CMV titre (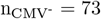, n_Low titre_ = 43, n_High titre_ = 27, *P* values derived from Kruskal-Wallis and post-hoc Dunn tests). Lower and upper box hinges represent the 25^th^ to 75th percentiles, the central line represents the median and whiskers extend to the highest and lowest values no greater than 1.5×IQR. **(b)** No association between CMV titre (a.u.ml^−1^) with HLA-A*02:01 pp65-reactive CD8^+^ T cells (n = 25, *rho* = −0.0065, *P* = 0.98, Spearman’s rank correlation test, *rho* denotes the Spearman *rho* and *P* value is from a two-sided *t* −test).**(c)** CD4^+^ T cells subsets according to titre (samples and boxplots as in a.). **(d)** Pre-treatment FOXP3^+^CD25^+^ T_regs_ are depleted in both CMV^+^_High titre_ and CMV^+^_Low titre_ groups relative to CMV^−^ patients (n = 98, 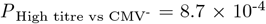, Est_adj_ = −1.3 and 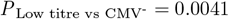, Est_adj_ = −0.94) when adjusting for treatment regimen, tumour type, age, and sex (results from a linear mixed model random effects analysis, centre point marks the estimate for the variable in association with FOXP3^+^CD25^+^ T_regs_ proportion of total CD4^+^ T cells, error bars mark the 95% CI, *P* values derived from ANOVA).

To validate the hypothesis that CMV^+^_High titre_ patients have an increased proportion of their CD8^+^ T cell repertoire occupied by cytotoxic T cells, we performed blinded flow cytometry, assessing major pretreatment subsets (n = 143). Interestingly, we found that the well-described CMV-related changes in T cell subset composition[27] were exacerbated in CMV^+^_High titre_ patients, with further skewing towards expansion of terminally differentiated effector memory cells (T_EMRA_) (*P* = 0.0020) and a reciprocal reduction in the proportion of T_Naïve_ cells (*P* = 0.0051) compared to CMV^+^_Low titre_ patients (Fig.4a). An expansion of T_EMRA_ is associated with increased effector function and is the hallmark of memory inflation, thus we postulated that the proportion of CMV-reactive T cells may directly relate to anti-CMV IgG titre. Using a smaller cohort of CMV^+^ patients who were genotyped for HLA-A*02:01 carriage (n = 25), we assessed the proportion of CD8^+^ T cells that were reactive to a tetramer loaded with the inflationary CMV pp65 epitope. Interestingly, despite the association with increased total CD8^+^ T_EMRA_ count, we found no correlation between titre and the proportion of CMV-reactive CD8^+^ T cells identified using this tetramer (*rho* = −0.0065, *P* = 0.98), suggesting that the non-tetramer-recognising T_EMRA_ cells were reactive to alternative inflationary CMV epitopes or that the increased expansion of the T_EMRA_ compartment with increasing anti-CMV IgG titre reflected inflation of non-CMV specific clones, reflecting a bystander effect of titre (Fig.4b).

Prior studies have suggested that CMV has profound immunomodulatory effects on CD4^+^ T cells, with a population of CD4^+^ T_EMRA_ being associated with CMV independently of age[28, 29]. To explore the relationship between anti-CMV IgG titre and CD4^+^ T cells, we assessed major subsets using the same flow cytometry panel described for CD8^+^ T cells. Here we noted a significant expansion of the effector memory (T_EM_) compartment in the CMV^+^_High titre_ compared to CMV^+^_Low titre_ patients (*P* = 0.0039); however, a consistent direction of effect was noted for T_Naïve_ and T_EMRA_ subsets to that observed in CD8^+^ T cells, suggesting that the enhanced cytotoxicity associated with an elevated titre is true in helper T cells. Finally, CMV^+^ patients were found to have reduced counts of T_regs_ (FOXP3^+^CD25^+^ regulatory T cells) in the periphery compared to CMV^−^ patients[12]. We therefore assessed the relationship between T_reg_ proportion of total CD4^+^ T cells (n = 98) in a multivariable analysis, correcting for tumour type, treatment intent, age, sex and experimental batch. We found that increasing age at onset of systemic treatment was associated with a slight increase in T_reg_ proportion (*P* = 0.034, Est_adj_ = 0.021) reinforcing the dichotomous effects of CMV from age-related immunosenescence[9, 30]. Interestingly however, both CMV^+^_Low titre_ and CMV^+^_High titre_ patients had a reduction of their T_reg_ compartment (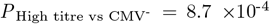, Est_adj_ = −1.3 and 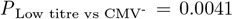, Est_adj_ = −0.94). Notably, we observed no difference in T_reg_ proportion within CMV^+^ patients (*P* = 0.37), indicating the effect of CMV on T_regs_ was influenced primarily by serostatus as opposed to total anti-CMV IgG titre.

## 3 Discussion

Here, in an updated extended cohort of 448 prospectively recruited patients receiving ICB for cancer, we replicate our previous observation from a less-mature subset of this cohort (n=308) of a protective effect of CMV serostatus against Grade 3+ irAEs. In this new analysis we further explore the relationship between the titre of CMV-reactive IgG antibodies and irAE risk. In seropositive patients, consistent with prior studies, we find the anti-CMV IgG titre remains stable over time [8, 16]. Additionally, anti-CMV IgG titre is independent of clinical covariates including sex, age and cancer type. Notably, while CMV^+^ patients have reduced risk of severe (Grade 3+) irAEs, we observe that those with above median anti-CMV IgG titres are further protected against severe irAEs. Although this protection was most notable in recipients of anti-CTLA-4 containing cICB, which has a markedly increased risk of irAEs, a strong trend towards protection was seen in the smaller group of sICB recipients (P=0.06), where our power to detect protection was much reduced. This suggests that high anti-CMV IgG titre is not only associated with limited priming of autoreactive immunity following anti-CTLA-4 but also limits non-specific anti-PD-1 responses[31].

Intriguingly, the protection attributable to high anti-CMV IgG titre can be shown to be a composite of organ-specific effects, with the CMV-related protection for non-colitis irAEs being confined to those with above median anti-CMV IgG titres. Conversely, the protective effect of CMV seropositivity against the development of colitis was independent of anti-CMV IgG, indicating colitis to be the most CMV-sensitive irAE. This observation provides further evidence for the risk factors of irAEs being organ-specific, inferring that pan-organ irAE studies have reduced sensitivity to detect both immunological and genetic associations. Moreover, our findings suggest that anti-CMV IgG titre reflects spatial segregation of CMV effects on immunity, in accordance with studies describing organ-specific T cell responses to CMV infection. The cause of this is unclear, although in murine models the titre of anti-CMV IgG relates to the magnitude of primary CMV infection [10, 32, 33].

Further analysis of the relationship between anti-CMV IgG titre and circulating T cells subsets demonstrated that CMV^+^_High titre_ patients had more pronounced cytotoxic responses compared to CMV^+^_Low titre_ patients across both CD4^+^ and CD8^+^ T cell compartments, reflecting greater repertoire occupancy of T_EMRA_ cells. This finding corroborates prior work demonstrating synergism between T cell and humoral anti-CMV immunity[34]. Interestingly however, we did not observe a significant association between titre and the proportion of CMV-reactive CD8 against the major tegument protein pp65, suggesting that humoral immunity is either linked to bystander T cell activation or that this association is restricted to CMV-specific CD4^+^ T cells, as previously suggested[34, 35]. Antigen-independent activation has been reported following several viral infections, specifically by means of IL-15 signaling, and this is an avenue that requires further exploration in the context of cancer immunotherapy[36, 37]. Finally, we find that although there is no significant difference in the proportion of circulating T_regs_ according to titre, CMV and age appear to have dichotomous effects on this compartment, consistent with prior murine and human studies highlighting an accumulation of T_regs_ with enhanced immunosuppressive capacity with age[38, 39]. Despite T_regs_ being a core component of peripheral tolerance, studies in colitis patients have found increased T_reg_ infiltration in the colon compared to controls[40]. Our work suggests that increasing circulating T_regs_ may be a marker of baseline inflammation in cancer and thus may aid in identifying patients who are at greater risk of developing toxicities, especially colitis.

Our study incorporates the largest analysis of the relationship between CMV status and cancer immunotherapy outcomes. Nonetheless, although the cohort is prospectively sampled, our findings are observational in nature. Whereas we previously noted no detectable CMV viraemias reflecting reactivation events in patients receiving ICB [12], we cannot exclude sub-threshold detection according to anti-CMV IgG titre. We are also limited currently to analysis of peripheral immunity and, whilst this is arguably the most informative tissue compartment from the perspective of biomarkers, further dissection of anti-CMV IgG titre on irAE development will require tissue directed work.

Development of irAEs forms the leading cause of patient morbidity during ICB treatment and may shape clinical decision processes. Therefore prospective, pre-treatment biomarkers are of high value. This work indicates that not only can CMV serostatus, a easily determined and affordable parameter, be used to guide propensity to Grade 3+ irAEs, but the anti-CMV IgG titre provides further clinical information with highly significant immune correlates. Specifically, patients who are seropositive for CMV are significantly less likely to develop colitis, irrespective of the anti-CMV IgG titre. Conversely, CMV protection against non-colitis irAEs is primarily conferred to those with an anti-CMV IgG titre of ≥ 180a.u.ml^−1^. CMV serology is a rapid baseline predictor of irAE risk that can be obtained from a liquid biopsy, allowing for easy scalability to standard-of-care testing in a clinical setting.

## 4 Methods

### 4.1 Ethics approval and consent to participate

Informed consent was given by all patients to donate samples to the Oxford Radcliffe Biobank (Oxford Centre for Histopathology Research ethical approval reference 19/SC/0173, project nos. 16/A019, 18/A064 and 19/A114) and grant access to their routine clinical data. There was no compensation for this consent.

### 4.2 Patients and clinical outcomes

Patients were recruited between 23 November 2015 and 19 July 2025. Patients eligible for ICB in the Oxford University Hospitals NHS Foundation Trust were recruited prospectively. These included patients with advanced or metastatic Melanoma, Renal Cell Carcinoma, Microsatellite instable Colorectal Cancer, Mesothelioma and Cutaneous Squamous Cell Carcinoma. Patients due to receive adjuvant ICB for Melanoma and Renal Cell Carcinoma were also eligible. Treatment regimens are listed in Supplementary Table 1. irAEs were assessed according to the National Cancer Institute’s Common Terminology Criteria for Adverse Events, version 5.0.

### 4.3 Sample collection

Up to 50ml of whole blood was collected from each patient in EDTA vacutainer tubes, immediately prior to the first dose of ICB. Plasma and PBMCs were separated after blood collection via density centrifugation using Ficoll-Paque (Cytiva). Anti-CMV IgG antibody titres were determined from plasma that was diluted 1:2 in HBSS on an Abbott Architect i2000 at the John Radcliffe Hospital, Oxford University Hospitals. Patients with an antibody titre of *<*4a.u.ml^−1^ were called CMV^−^. For CMV+ patients saturating values (*>*250a.u.ml^−1^) were called as 250a.u.ml^1^, median value = 89.4a.u.ml^−1^ (mean = 109.3a.u.ml^−1^, IQR: 54.4-158.8a.u.ml^−1^). CD8^+^ T cells were isolated from PBMC for bulk RNA-sequencing using Miltenyi Biotech magnetic separation via positive selection as per the manufacturer’s instructions. PBMCs were counted and stored in liquid nitrogen at 5-10 × 10^6^ cells/vial.

### 4.4 Nucleic acid extraction

Positively selected CD8^+^ T cells (bulk RNA-sequencing) and PBMCs (genotyping) were spun down at 4°C and resuspended in 350µl of RLTplusbuffer (Qiagen) supplemented with 2% DTT and stored at −80°C for DNA/RNA extraction. Samples were homogenized using the QIAshredder kit (Qiagen). DNA/RNA was extracted using the AllPrep Universal DNA/RNA/miRNA Kit (Qiagen). DNase I was used during the RNA extraction protocol to minimize DNA contamination. RNA was eluted into RNase-free water and quantified via Qubit analysis. Samples were stored at −80°C until library preparation.

### 4.5 Bulk RNA-sequencing

RNA was thawed on ice and messenger RNA was isolated using the NEBNext Poly(A) mRNA Magnetic Isolation Module Kit. Library preparation was performed using NEBNext Ultra II Directional RNA Library Prep Kit for Illumina. mRNA was then sequenced using 150-base pair (bp)-end sequencing using an Illumina NovaSeq 6000/ NovaSeq XPlus or 75bp-end sequencing on an Illumina HiSeq-4000, at Genewiz and the Oxford Genome Centre, respectively. Reads were aligned to hg38 build of the human genome using HISAT2 and read count data was generated using HTSeq[41, 42]. High quality reads were chosen based on MAPQ score provided by bamtools[43]. Marking and removal of duplicated reads was performed using picard while samtools was used to calculate statistics[44]. DESeq2 (v.1.48.1) was used to perform differential expression analysis between CMV^+^_Low titre_ and CMV^+^_High titre_ patients’ pre-treatment samples [45]. Age, sex, tumour type, treatment intent, and the first principal component of all genes were adjusted for in the design formula and only genes with a mean expression *>*10 were included in the analysis. Normalised expression of *IFNG, KLRD1, TBX21* and *ZNF683* was also determined using DESeq2[45].

### 4.6 Flow Cytometry

Cryopreserved patient PBMC samples were thawed at 37°C and washed with HBSS. 1 × 10^6^ PBMCs were plated, and viability staining was performed with eBioscience Fixable Viability Dye eFluor780 (Invitrogen, cat. no. 65-0865-14) or Fixability Viability Stain 440UV (BD Biosciences) for 30min at 4°C. PBMCs were washed with HBSS supplemented with 5% FCS and BD Horizon Brilliant Stain Buffer (BD Biosciences, 563794). Staining for T cell surface markers was performed for 30min at 4°C using antibodies directed against markers listed in Supplementary Table 4. PBMCs were washed once in cell staining buffer (BioLegend, cat. no. 420201) and incubated with Streptavidin (BV785, BioLegend cat. no. 405249) for 30min at 4°C for IgG4 staining. Following surface staining, PBMCs were permeabilized with Foxp3/Transcription Factor Staining Buffer Set (Invitrogen, cat. no. 00-5523-00) for intra-nuclear staining or Transcription Factor Buffer Set (BD Biosciences, cat. no. 562574) for cytoplasmic staining, for 30min, and incubated with intracellular staining antibodies listed in Supplementary Table 4. PBMCs were resuspended in 2% paraformaldehyde and acquisition was performed on a FACSSymphony A5 Cell Analyzer (BD Biosciences) or a Fortessa X-20. The CMV-tetramer panel was acquired on an Attune NxT. Data was analyzed using FlowJo v.10.7.1 (BD Biosciences).

### 4.7 HLA-typing

Genotyping was performed using the Illumina Global Screening Array 24 version 3. The vcf files for chromosome 6 were inputted into the Michigan Imputation Server Genotype Imputation HLA (Minimac4) 1.5.8 pipeline against the Multi-ethnic HLA reference panel. Results were filtered by Class I and II genes.

### 4.8 Statistical analysis

Statistical analysis was performed using R v.4.5.1. Cumulative incidence and competing risk regression was performed using tidycmprsk (v1.1.0) to account for guarantee-time bias. Univariate estimates were calculated for time to irAE development using a semi-competing risks model, where death was the competing risk. Significance testing was performed using a Gray’s test. Competing risk regression was used to calculate the Fine-Gray sub-distribution hazard for association of CMV titre and each irAE. Distributions were compared using either a Wilcoxon signed-rank test, for paired data, or a Wilcoxon rank sum test for unpaired data. For multiple comparisons, a Kruskal-Wallis test was used followed by a post-hoc Dunn test. Correlation analysis was performed using a Spearman’s rank correlation test. Estimates for FOXP3^+^CD25^+^T_reg_ proportions were obtained using a linear mixed effects model where batch was a random effect. All multivariable analysis included age, sex, treatment regimen, and tumour type as covariates unless stated otherwise. Plots were generated using ggplot2 (v3.5.2). For all data, \**P<*0.05, \*\**P<*0.01, \*\*\**P<*0.001, \*\*\*\**P<*0.0001.

## Supporting information

Supplementary Tables, Age grouped

## 5 Data availability

Raw sequencing data corresponding to anonymized patients will be made available for download from EGA (XXXX) via a Data Access Arrangement between applicants and the University of Oxford, 6 months from the date of publication—these data will be available for a minimum of 2 years. Requests should be addressed to and will be processed within the university but should take *<*6 weeks to action. Normalized gene expression matrices with associated minimal anonymized patient information (required to perform analyses to recreate the figures in the paper) is available via Oxford Research Archive (https://doi.org/10.5287/ora-5jy9jpmaq).

## 6 Code availability

Code to perform all analyses and reproduce main and extended figures can be found on the Fairfax lab GitHub (https://github.com/fairfaxlab).

## 7 Acknowledgements

We are grateful to all patients who contributed samples and participated in the study without compensation. We thank all the staff of the Day Treatment Unit, Oxford Cancer Centre and the Brodey Centre at the Horton General Hospital. We are grateful to all the staff of the Oxford University Hospitals NHS Foundation Trust haematology, microbiology and biochemistry laboratories and thank T. James for his facilitation, as well as the staff of the Oxford Radcliffe Biobank and Churchill Hospital Sample Handling Lab.

## 8 Author contributions

B.P.F. conceptualized and oversaw the study. B.P.F. & G.M. were responsible for the methodology. G.M., M.L., S.M., O.T., D.M., A.F., A.H., C.A.T performed investigations. G.M. & S.M. performed visualizations. B.P.F. was responsible for funding acquisition. B.P.F., M.L., M.R.M. and S.M. were responsible for project administration. B.P.F. supervised the study. B.P.F., M.L., M.R.M. were responsible for resources. G.M. performed the formal analysis. M.L., S.M., G.N. and S.S. were responsible for data curation. G.M., S.S., and G.N. were responsible for software. B.P.F. and G.M. wrote the original draft of the manuscript. B.P.F., G.M., M.R.M. and P.K. reviewed and edited the manuscript.

## 11

**Extended Data Figure 1:**
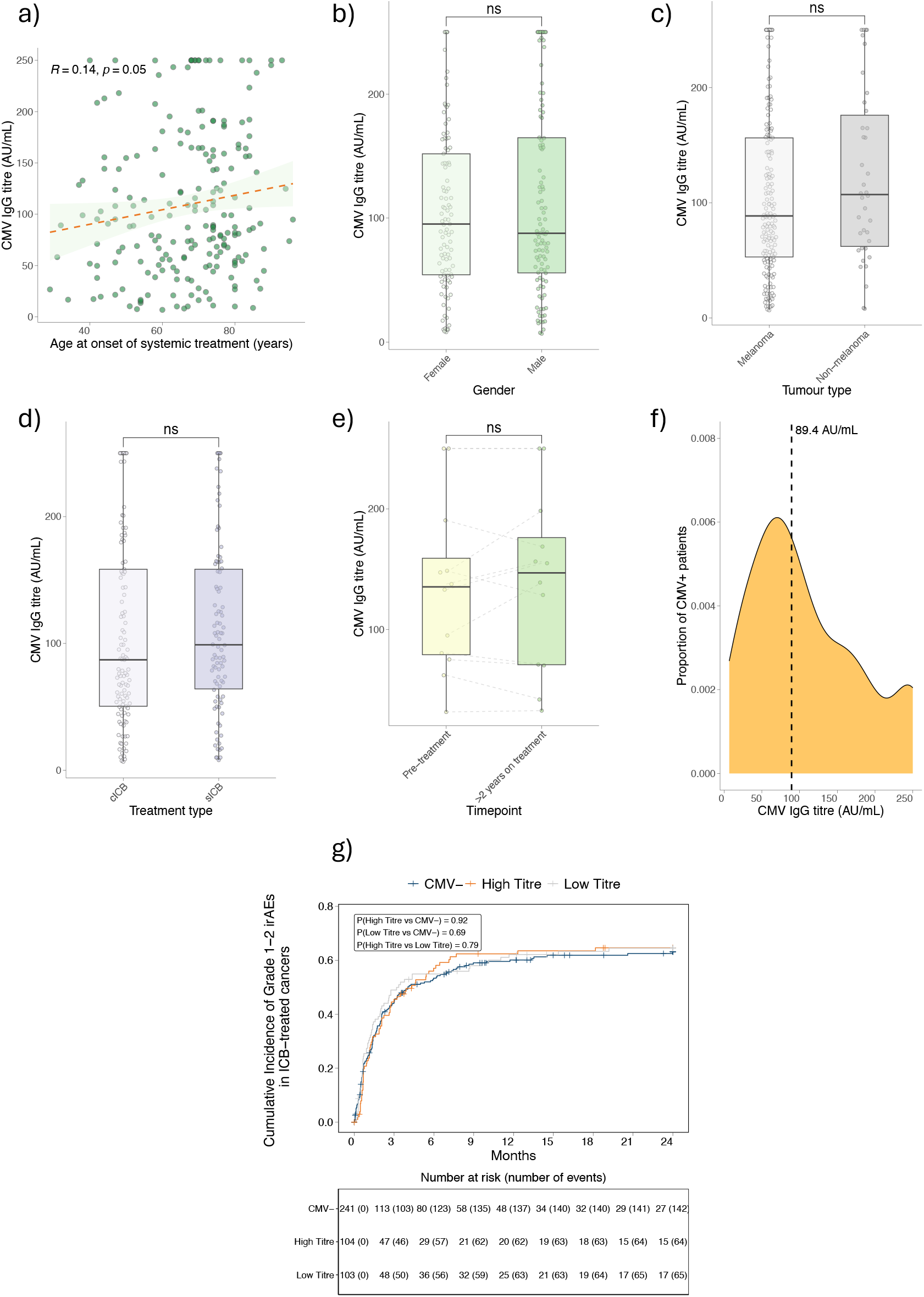
Clinical correlates of anti-CMV IgG titre. **(a)** Correlation of anti-CMV IgG titre (a.u.ml^−1^) and age at onset of ICB treatment (n = 207, Spearman’s rank correlation test, R denotes the Spearman *rho* and *P* value is from a two-sided *t* −test).**(b)** Anti-CMV IgG titre (a.u.ml^−1^) according to biological sex (n_Male_ = 106, n_Female_ = 101, *P* = 0.71). *P* values result from the application of a Wilcoxon rank sum test; lower and upper box hinges represent the 25th to 75th percentiles, the central line represents the median and whiskers extend to the highest and lowest values no greater than 1.5×IQR. **(c)** anti-CMV IgG titre (a.u.ml^−1^) according to tumour type (n_Melanoma_ = 169, n_Non-melanoma_ = 38, *P* = 0.2, Wilcoxon rank sum test, boxplot as in b.). **(d)** anti-CMV IgG titre (a.u.ml^−1^) according to treatment regimen (n_cICB_ = 113, n_sICB_= 94, *P* = 0.41, Wilcoxon rank sum test, boxplot as in b.). **(e)** anti-CMV IgG titre (a.u.ml^−1^) at pre-treatment and late timepoints (*>*2y on treatment) (n = 12 patients, *P* = 0.68, Wilcoxon signed-rank test, boxplot as in b.). **(f)** Distribution of anti-CMV IgG titres (a.u.ml^−1^) in seropositive cohort (n = 207) with median titre (89.4a.u.ml^−1^ indicated). **(g)** Cumulative incidence of Grade 1-2 irAEs according to CMV titre (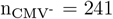, n_Low titre_ = 103, n_High titre_ = 104, two-sided log-rank test)

**Table 1:** Cohort matching table for Grade 3+ irAE development.

| Characteristic | Grade 3+ irAEs |  | p-value<sup>2</sup> |
| --- | --- | --- | --- |
|  | Developed<sup>1</sup><br>N = 172 | Not developed<sup>1</sup><br>N = 276 |  |
| age | 65 (53, 73) | 70 (59, 77) | <0.001 |
| sex |  |  | 0.8 |
| Female | 72 (39%) | 113 (61%) |  |
| Male | 100 (38%) | 163 (62%) |  |
| cancer |  |  | <0.001 |
| Colon | 2 (13%) | 14 (88%) |  |
| cSCC | 1 (9.1%) | 10 (91%) |  |
| Melanoma | 157 (43%) | 208 (57%) |  |
| Mesothelioma | 8 (32%) | 17 (68%) |  |
| RCC | 4 (13%) | 27 (87%) |  |
| Treatment |  |  | <0.001 |
| Adjuvant | 8 (13%) | 53 (87%) |  |
| cICB | 141 (51%) | 137 (49%) |  |
| sICB | 23 (21%) | 86 (79%) |  |

<sup>1</sup> Median (Q1, Q3); n (%)
<sup>2</sup> Wilcoxon rank sum test; Pearson's Chi-squared test; Fisher's exact test

**Table 2:** Breakdown of organ-specific Grade 3+ irAEs according to CMV titre.

| Characteristic | CMV-<br>N = 156<sup>1</sup> | High Titre<br>N = 25<sup>1</sup> | Low Titre<br>N = 62<sup>1</sup> | Events in all patients |
| --- | --- | --- | --- | --- |
| organ |  |  |  |  |
| Cardiac | 2 (33%) | 0 (0%) | 4 (67%) | 6 / 448 |
| Colitis | 57 (78%) | 8 (11%) | 8 (11%) | 73 / 448 |
| Dermatitis | 5 (38%) | 2 (15%) | 6 (46%) | 13 / 448 |
| Endocrine | 19 (61%) | 6 (19%) | 6 (19%) | 31 / 448 |
| Gastritis | 8 (57%) | 1 (7.1%) | 5 (36%) | 14 / 448 |
| Hepatitis | 26 (57%) | 6 (13%) | 14 (30%) | 46 / 448 |
| Nephritis | 3 (38%) | 0 (0%) | 5 (63%) | 8 / 448 |
| Neurological | 5 (50%) | 2 (20%) | 3 (30%) | 10 / 448 |
| Other | 7 (70%) | 0 (0%) | 3 (30%) | 10 / 448 |
| Pneumonitis | 8 (89%) | 0 (0%) | 1 (11%) | 9 / 448 |
| Rheumatological | 16 (70%) | 0 (0%) | 7 (30%) | 23 / 448 |
| <sup>1</sup> n (%) |  |  |  |  |

## 12 Supplementary information

### 12.1 Supplementary Tables

- **Supplementary Table 1** Treatment regimens in the OxCITE cohort
- **Supplementary Table 2** Differentially expressed CD8^+^ T-cell genes between CMV^+^_Low titre_ and CMV^−^ patients
- **Supplementary Table 3** Differentially expressed CD8^+^ T-cell genes between CMV^+^_High titre_ and CMV^−^ patients
- **Supplementary Table 4** Flow cytometry panels
- **Supplementary Table 5** Cohort missingness table

